# Estimating the burden of United States workers exposed to infection or disease: a key factor in containing risk of COVID-19 infection

**DOI:** 10.1101/2020.03.02.20030288

**Authors:** Marissa G. Baker, Trevor K. Peckham, Noah S. Seixas

## Abstract

**Introduction:** With the global spread of COVID-19, there is a compelling public health interest in quantifying who is at increased risk of disease. Occupational characteristics, such as interfacing with the public and being in close quarters with other workers, not only put workers at high risk for disease, but also make them a nexus of disease transmission to the community. This can further be exacerbated through presenteeism, the term used to describe the act of coming to work despite being symptomatic for disease. Understanding which occupational groups are exposed to infection and disease in the workplace can help to inform public health risk response and management for COVID-19, and subsequent infectious disease outbreaks.

**Methods:** To estimate the burden of United States workers exposed to infection and disease in the workplace, national employment data (by Standard Occupational Classification) maintained by the Bureau of Labor Statistics (BLS) was merged with BLS O*NET survey data, which ranks occupations with particular physical, ergonomic, and structural exposures. For this analysis, occupations reporting exposure to infection or disease more than once a month was the focus.

**Results:** Based on our analyses, approximately 10% (14.4 M) of United States workers are employed in occupations where exposure to disease or infection occurs at least once per week. Approximately 18.4% (26.7 M) of all United States workers are employed in occupations where exposure to disease or infection occurs at least once per month. While the majority of exposed workers are employed in healthcare sectors, other occupational sectors also have high proportions of exposed workers. These include protective service occupations (e.g. police officers, correctional officers, firefighters), office and administrative support occupations (e.g. couriers and messengers, patient service representatives), education occupations (e.g. preschool and daycare teachers), community and social services occupations (community health workers, social workers, counselors), and even construction and extraction occupations (e.g. plumbers, septic tank installers, elevator repair).

**Conclusions:** The large number of persons employed in a wide variety of occupations with frequent exposure to infection and disease underscore the importance of all workplaces developing risk response plans for COVID-19. This work also serves as an important reminder that the workplace is a key locus for public health interventions, which could protect both workers and the communities they serve.

## Introduction

As COVID-19 spreads globally, there is public health importance in characterizing the role of the workplace in disease transmission, given the amount of time people spend at work, and the variety of work tasks that could promote the spread of infectious disease (e.g., interfacing with customers, patients, and co-workers; preparing food). Presenteeism, reporting to work despite being symptomatic for disease, commonly occurs in the workplace, and can contribute to the transmission of infectious disease, and potentially to the spread of epidemics or pandemics (1,2). One analysis examined the role of workplace transmission in the 2009 H1N1 pandemic, estimating that about 8 million employees in the United States worked while infected, and that these workers may have caused the infection of as many as 7 million of their co-workers (3).

The United States Centers for Disease Control and Prevention (CDC) has published interim guidance for businesses and employers to plan and respond to COVID-19, including guidance for actively encouraging employees to stay home, separating sick employees, encouraging respiratory and hand hygiene for all employees, performing environmental cleaning, and providing specific advice for employees who need to travel (4). However, the best guidance for workplaces will be geared to be specific to a particular occupational sector that could be exposed to COVID-19. It is known that those working in healthcare settings are at increased risk for exposure to infectious diseases such as COVID-19, but may also have better infectious disease protection plans than other occupational settings. In 2007, Rebmann et al. (5) surveyed 633 Infection and Prevention Control Professionals that work at hospitals to understand the infection control policies at hospitals across the country. Rebmann et al. found that most hospitals employ infection control professionals, have a health care worker prioritization plan for vaccines or antivirals, and have plans in place to stockpile N95 respirators and medication, all of which could help control exposure for workers in the hospital. While important, these measures may be inadequate for the effective prevention of infection for such high risk occupations (6). Other occupational groups which may have more sporadic exposure to infection or disease may not have the same level of planning, or even think that an infection disease control plan is warranted for their workplace. Therefore, it is important to understand which occupations could be exposed to infection or disease in order to encourage these occupational sectors to develop and implement an infectious disease response plan for disease outbreaks such as COVID-19.

While several groups have broadly characterized the burden of occupational injury or illness (7–9) fewer groups have characterized the burden of occupational exposure, and rarely do these burden of exposure estimates include exposure to biologic agents, disease, or infection in the workplace (10,11). Montano (2014) systematically reviewed which occupational groups in Europe have risk of exposure to biological agents, finding workers in healthcare, biotechnology, agriculture, elementary education, craft work, operators, and the armed forces were exposed to various pathogens; the number of workers exposed and frequency of exposure was not reported (12). Focusing on outcome as opposed to exposure, Anderson et al. characterized the distribution of influenza-like illness (ILI) by occupation in Washington state, finding janitors, cleaners, and janitors more likely to catch ILI (13).

Understanding the burden of occupational exposure to infection and disease, including how many workers are potentially exposed and what occupations they work in, allows for upstream prevention measures, such as workplaces developing appropriate infectious disease response plans, integrating infectious disease trainings into other workplace trainings, and beginning to develop workplace policies that can support a workforce potentially exposed to COVID-19. This will help reduce the transmission of infectious disease from and within the workplace.

Previously, state-level employment data were utilized to estimate the number of workers exposed to a host of occupational exposures in Federal Region X (WA, OR, ID, AK), spanning chemical, physical, ergonomic, and psychosocial hazards (14). Here, utilizing the same data analysis methods as previously detailed in Doubleday et al., the number of workers across the United States exposed to disease or infection at work more than once a month is estimated. We believe this analysis is valuable for informing risk assessments and protective actions that occupational sectors can take during infectious disease outbreaks, such as COVID-19.

## Methods

Two sources of data were utilized for this analysis, and are detailed below.

United States employment data was obtained from the Bureau of Labor Statistics (BLS) Occupational Employment Statistics database (15). The most current employment data at the time of analysis was from May 2018, and is organized by 2010 Standard Occupational Classification codes (2010 SOC). SOC codes are hierarchical, ranging from two-digits (Major Group Code) to six digits (Detailed Occupation Code), with the six-digit codes being the most detailed (16).

To estimate exposure to disease and infection in the workplace, we used data within the O*NET database. O*NET is a job characterization tool, with rich information on tasks performed, skills needed, and job characteristics for different occupations, in order to inform job seekers or researchers (17). As nearly 600 six-digit SOC occupations are updated each year, the entire O*NET database is completely refreshed every few years (18). Between 2001 and 2011, nearly 160,000 employees from 125,000 workplaces had responded to O*NET questionnaires, and no SOC codes are missing from the data (19).

To characterize risk of workplace exposure to infectious disease, we used the following O*NET question: “How often does this job require exposure to disease/infections?” Respondents could select from the following multiple choice answers: Never; Once a year or more but not every month; Once a month or more but not every week; Once a week or more but not every day; Every day (20). Within O*NET, these data are converted to a 0-100 score, representing weighted-average frequency or intensity of the metric for each SOC code. For this analysis, occupations were retained that had a score of 50-100, representing exposure to disease/infection more than once a month. SOC codes were merged with the national employment data to calculate the total number of workers employed in the occupations with exposure to disease/infection at more than once a month.

All data analysis was conducted using the statistical software package R version 3.6.3.

## Results

As of May 2018, there were a total of 144.7 million persons employed in the United States in employer-employee arrangements counted by BLS. Of these 144.7 million workers, an estimated 18.4% (26,669,810) were employed in occupations where exposure to disease or infection occurs more than once a month. As of May 2018, 10% (14,425,070) of the United States workforce was employed in occupations where exposure to disease or infection occurs at least once a week. Table 1 summarizes the major occupational sectors (two-digit SOC) that have detailed occupations (six-digit SOC) exposed to infection or disease more than once a month, and the number of workers this represents. Both Healthcare Practitioner and Technical Occupations, and Healthcare Support Occupations have more than 90% of workers exposed more than once a month, and more than 75% of workers exposed more than once a week. Other notable major occupation groups with high proportion of exposure are Protective Service Occupations (52% exposed more than once a month, including police officers, firefighters, transportation security screeners), Personal Care and Service Occupations (52% exposed more than once a month, including childcare workers, nannies, personal care aides), and Production Occupations (60% exposed more than once a month, including laundry and dry cleaning workers, wastewater treatment operators, dental technicians). The 16% of office and administrative support occupations with exposure to disease or infection more than once a month are patient representatives, couriers and messengers, and medical secretaries.

**Table 1:** Number and percent of workers exposed to infection or disease more than one time per month, and more than one time per week, by major (2-digit) standard occupational classification code (SOC). Major SOCs which did not have any occupational groups exposed more than one time per month are not included in the above table.

|  |  |  | Exposed > 1 time/month |  | Exposed > 1 time/week |  |
| --- | --- | --- | --- | --- | --- | --- |
| 2-digit SOC |  | total in SOC | # | % | # | % |
| 11 | Management | 7,616,650 | 59,050 | 0.8% | -- | -- |
| 13 | Business and Financial Operations | 7,721,300 | 300,900 | 3.9% | 300,900 | 3.9% |
| 15 | Computer and Mathematical | 4,384,300 | 587,970 | 13.4% | -- | -- |
| 19 | Life, Physical, and Social Science | 1,171,910 | 159,970 | 13.7% | 20,030 | 1.7% |
| 21 | Community and Social Services | 2,171,820 | 704,280 | 32.4% | 168,190 | 7.7% |
| 25 | Education, Training, and Library | 8,779,780 | 2,048,070 | 23.3% | -- | -- |
| 27 | Arts, Design, Entertainment, Sports, and Media | 1,951,170 | 57,140 | 2.9% | -- | -- |
| 29 | Healthcare Practitioners and Technical | 8,646,730 | 7,911,430 | 91.5% | 6,728,420 | 77.8% |
| 31 | Healthcare Support | 4,117,450 | 3,958,560 | 96.1% | 3,160,890 | 76.8% |
| 33 | Protective Service | 3,437,410 | 1,789,490 | 52.1% | 1,026,660 | 29.9% |
| 37 | Building and Grounds Cleaning and Maintenance | 4,421,980 | 924,290 | 20.9% | -- | -- |
| 39 | Personal Care and Service | 5,451,330 | 2,841,730 | 52.1% | 29,810 | 0.5% |
| 43 | Office and Administrative Support | 21,828,990 | 3,532,530 | 16.2% | 2,871,400 | 13.2% |
| 47 | Construction and Extraction | 5,962,640 | 491,990 | 8.3% | -- | -- |
| 51 | Production | 622,790 | 371,480 | 59.6% | -- | -- |
| 53 | Transportation and Material Moving | 10,244,260 | 930,930 | 9.1% | 118,770 | 1.2% |
| All SOCs (including those not listed above) |  | 144,944,620 | 26,669,810 | 18.4% | 14,425,070 | 10.0% |

## Discussion

During an infectious disease outbreak, the workplace can play an important role in both spreading the disease (21,22) and helping to stop the spread of disease through workplace practices and policies (23,24). Understanding the wide range of occupations that could be exposed to infection or disease due to work activities is important for planning risk management and communication to workers, in addition to prioritizing workplace response plans. This brief report estimates occupational groups that face increased risk of potential exposure to an infection or disease; however, estimates of the number of workers who fall ill due to such exposures are not possible in this analysis. However, a primary goal of public health, especially in the face of a global pandemic, is to prevent the spread of disease. Therefore, understanding who is potentially exposed is an important first step in being able to enact risk reduction strategies prior to disease transmission occurring, and illness manifesting. Therefore, the results reported here have important public health implications.

Some limitations must be noted. O*NET data were generated from self-reported subjective questionnaires and therefore are subject to bias and misclassification. Respondents may not realize they are exposed to infection or disease at work unless they are in a workplace where these hazards are communicated to them and protective equipment is provided (e.g., healthcare sectors) leading to potential differential misclassification across occupational groups. Additionally, information from the O*NET database is applied at the occupation-level, and therefore does not account for within-job exposure variation (25).

BLS employment estimates do not capture data on all workers in the United States, including self-employed, undocumented, continent, and domestic workers. These workers may be uniquely at risk to exposures to work because due to limited ability to take time off if they or a family member is ill (26). In Sweden and Norway, higher rates of presenteeism (coming to work when sick) were found among low-income and immigrant workers (27).

Access to paid leave, which could ameliorate the financial burden of staying home while sick, varies substantially by occupation, industry, employer, location, and worker sociodemographic profile (e.g., race/ethnicity) (28,29). Workers without access to paid leave have higher rates of presenteeism, and are less likely to receive preventative health services such as getting flu shots (30). Occupational sector also influences rates of presenteeism, with studies from various countries showing higher rates of presenteeism among workers in healthcare, public service, and educational sectors, as these essential services often do not have substitute workers available (31–33). Indeed, a recent systematic review identified occupation type as one of the strongest predictors of presenteeism (1). As many of these sectors are already at risk of exposure to disease due to work activities, it is important that disease response plans for these sectors include not only control methods to reduce exposures at work, but also contingency plans to ensure sick workers do not come back to work with disease. This could be accomplished through cross-training, providing extra paid sick leave during this time, ensuring flexible working conditions, and ensuring substitute workers are identified to fill in if essential workers fall ill.

In conclusion, our analysis shows that a large proportion of the United States workforce, across a variety of occupational sectors, are exposed to disease or infection at work more than once a month. These are workers that public health should consider especially at risk to exposure to COVID-19. However, it should be noted that there are many other workers that could also be uniquely at risk to exposure to COVID-19, or encourage the spread of COVID-19, such as workers who are not given access to flexible working, workers who do not feel they can take sick time if they or a family member is sick, workers who do not have access to paid sick leave, or workers that perform essential services and do not have access to substitute workers. Work presented here underscores the importance of all workplaces developing sector-specific response plans to keep employees safe, halt the transmission of disease in the workplace, and ensure sick workers do not have to come to work. It also serves as a reminder that the workplace is an important locus for public health interventions, that can affect both the workers and the community.

## Data Availability

All data is publicly available online, links to this are referenced in the submission.

https://www.bls.gov/oes/home.htm

https://www.onetonline.org

## Acknowledgments

The authors gratefully acknowledge Annie Doubleday for developing the R code that supported this analysis.

